# Population level risk stratification of hypertensive patients by predictive modeling

**DOI:** 10.1101/2021.04.27.21256198

**Authors:** Sricharan Bandhakavi, Zhipeng Liu, Sunil Karigowda, Jasmine M. McCammon, Farbod Rahmanian, Heather M. Lavoie

## Abstract

**Objective:** We recently reported that hypertension (HTN) patients having <u>at l</u>east <u>three r</u>ounds <u>o</u>f distinct treatment <u>o</u>ptions (*atl_three_roto*) in a 12-month window have elevated risk of next-year complications. However, early identification of these “challenge to treat” patients is non-trivial and “drivers” of complications in these *vs* remaining HTN patients are not fully defined. To address these challenges/gaps, we present predictive models for preceding outcomes, delineate their “drivers”, and highlight value of their integration for population level risk stratification/management of HTN patients.

**Materials and Methods:** 2.47 million HTN patients enrolled through 2015-2016 were selected from a nation-wide commercial claims database. Features associated with their treatment patterns, comedications, and comorbidities were extracted for 2015 and used to model/predict 2016 outcomes of *atl_three_roto* status and/or HTN complications. Logistic regression-derived odds-ratios were used to delineate drivers of each outcome.

**Results:** Prior year treatment patterns, specific hypertension drugs (anti-hypertensives, calcium channel blockers, beta blockers), and congestive heart failure most increased future odds of *atl_three_roto* status. Regardless of prior year *atl_three_roto* status, specific comorbidities (renal disease, congestive heart failure, myocardial infarction, vascular disease, diabetes with chronic complications) and comedications (beta blockers, cardiac agents, anti-lipidemics) most increased future odds of HTN complications. Proof-of-concept analysis with an independent dataset demonstrated that integrating these model predictions/drivers thereof can be leveraged for risk stratification/management of HTN patients.

**Discussion:** Integrating predictions and their “drivers” from above models supports early identification and targeted management of “at-risk” HTN patients.

**Conclusion:** We have developed a predictive modeling based approach for risk stratification and management of HTN patients.

## BACKGROUND AND SIGNIFICANCE

Among chronic diseases in the United States, hypertension (HTN) remains one of the most prevalent and challenging to control [1]. Even among those being treated for HTN, >25% of patients do not have their blood pressure under control (i.e., within systolic/diastolic blood pressure limits of 130/80) [1]. The CDC has recently reported even higher prevalence of uncontrolled HTN and proposed that the majority of these HTN patients would benefit from treatment “adjustments” such as increased dosage of their current medication choice(s) or additional medications [2]. However, in the absence of supporting clinical information for all patients and the known association of (especially when uncontrolled) HTN with various other complications [3-5], there is a need to develop targeted population-based strategies for early identification/management of “high-risk” HTN patients (*e*.*g*. via use of administrative claims data).

To help with early identification of “high-risk” HTN patients from administrative claims, researchers have developed models for predicting HTN patients at risk of future (HTN-related) complications [6 7], including those that are grouped by severity [8]. Multiple groups have also performed treatment pathways/outlier analyses at scale to characterize prescription patterns among new HTN patients [9 10], compare various first line treatments for efficacy/safety [11], and identify future risk-associated treatment patterns among new *vs* all HTN patients [12]. Finally, evidence-based guidelines combined with electronic health records can be leveraged to identify putative gaps-in-care for HTN patients for future risk reduction [13-15].

We recently analyzed treatment patterns among > 4.1 million HTN patients from a nationwide administrative claims’ database for HTN-specific treatment (Rx) choices, Rx count, and distinct rounds <u>o</u>f treatment <u>o</u>ptions (<u>ROTO</u>), respectively [12]. Distinct ROTOs were identified (for each patient) by the first pick-up date(s) of new HTN medication(s) not previously seen within a calendar year/12-month observation period. This approach enabled identification of patients requiring multiple changes/additions to their HTN treatment(s). Based on further risk assessment, HTN patients having <u>at l</u>east <u>three r</u>ounds <u>o</u>f distinct treatment <u>o</u>ptions (henceforth referred to as *atl_three_roto* patients *i*.*e*., those with ROTO count ≥3; account for < 10% of all HTN patients) in a 12-month window had elevated odds of next year HTN complications (3.8-fold higher) vs those maintained with single/initial round of treatment options (henceforth referred to as *one_roto* patients, *i*.*e*., patients with ROTO count = 1 signifying no subsequent Rx changes/additions within calendar year; account for >60% of all HTN patients) [12]. Potential reasons for the multiple Rx changes/additions in *atl_three_roto* patients include inadequate responsiveness to initial/previous HTN medications, incorrect initial dosages, undesirable side-effects, and/or effects on other co-morbidities. Thus, *atl_three_roto* patients may be viewed as a unique “challenge to treat for HTN” sub-group and predictive approaches are needed for their early identification and targeted intervention.

In this study, we present a “challenge to treat for HTN” predictive model that can be used to predict which members will have <u>at l</u>east <u>three r</u>ounds <u>o</u>f treatment options (future *atl_three_roto* status) in the next 12-months. In addition, to better support targeted intervention strategies, we identified comedications, comorbidities, and treatment patterns associated with future HTN complications in *atl_three_roto* vs remaining HTN patients. Finally, we integrated these drivers/predictions to demonstrate their utility for risk stratification and management of HTN patients.

## MATERIALS AND METHODS

### Study design and cohort

We used commercially licensed, nationwide administrative claims data for the period between January 01, 2015 to December 31, 2016 for developing the predictive models described this study. Through the rest of this manuscript we refer to this data source as the CCAE (<u>C</u>ommercial <u>C</u>laims <u>a</u>nd <u>E</u>ncounters) database. Members who were enrolled for both medical and pharmacy benefits during the study period were processed to generate a study cohort of 2.47 million patients with HTN and subsequently used for predictive models described in next section. As an independent dataset (referred to as IND_DATA in rest of manuscript), commercially licensed administrative claims from an independent payer not in the CCAE database were processed identically to generate a cohort of 153,003 HTN patients for validating model performance/overlap between model predictions. In both cohorts, all patients were ≤ 65 years old.

For the study cohorts, members with HTN were identified based on their satisfying one of two criteria: member had at least 1 Rx claim for HTN-related medication(s) OR member had HTN-related diagnosis codes on distinct medical claims on at least two different dates. For identification of HTN-related medications, following <u>a</u>natomical therapeutic <u>c</u>lassification (ATC) system codes were used: C02, C03 except C03C, C04AB, C07, C08, C09 based on an independent report[16]. HTN-diagnosis codes were as previously described [8]. SQL and Python scripts were used for identification of study cohort, feature generation, predictive modeling analyses, and characterization of model outputs/predictions described in further detail below.

### Challenge to treat for HTN predictive model

Based on better performance when compared against a random forest model (tested initially as an alternative modeling algorithm; data not shown), a logistic regression model was used to predict those HTN patients in our study cohort who will be “challenging to treat for HTN” in the following 12-months. Patients who had <u>a</u>t least <u>three</u> distinct rounds <u>o</u>f HTN treatment <u>o</u>ptions in 2016 administrative claims data were labeled/defined as *atl_three_roto*/ “challenge to treat for HTN” patients (outcome variable). For this labeling, distinct rounds <u>o</u>f treatment <u>o</u>ptions (for HTN drugs) were counted for each member in study cohort for 2016 as described previously[12]. In brief, distinct round of treatment option(s) (ROTOs) were identified by the first pick-up date(s) of new HTN medication(s) – based on distinct 4^th^ level ATC code(s) - not seen previously for that member during the 2016 calendar year.

Input features (see **Supplementary Table S1**) for this model were calculated from 2015 administrative claims data (*i*.*e*., prior 12-months as all members were continually enrolled in 2015 and 2016) and included demographic features, <u>C</u>harlson <u>C</u>omorbidity Index (CCI)-based disease diagnosis groups (ICD-9/10 codes) [17], distinct medication types (identified by their ATC codes) and representing chronic conditions featured in <u>c</u>hronic <u>d</u>isease score (CDS) [18 19], and Rx/Dx claim-based patterns (distinct HTN-ROTO counts, distinct HTN-Rx counts, distinct Dx dates with HTN claims). Numeric features (age) were scaled between 0-1 and all categorical features were one-hot encoded. Most important features driving outcome variable (“challenge to treat for HTN” i.e., future *atl_three_roto* status in 2016) were identified by logistic regression-derived odds-ratios.

### Drivers of future HTN complications

After stratifying above study cohort by *atl_three_roto* status in 2015, logistic regression models were generated to predict among *atl_three_roto* vs *not_atl_three_roto* patients in 2015 who will develop “any stage” HTN complications (outcome variable) in 2016. Patients with “any stage” HTN complications were identified from 2016 administrative claims with ICD-9/10 diagnostic codes corresponding to increasing levels of severity (stage 1, stage 2, or stage 3 complications) across different target organ systems, as described in a previous report [8]. Input features (see **Supplementary table S2**) for the two predictive models were calculated using 2015 administrative claims data and included: demographic features, disease diagnosis groups, medication types, and 3 categorical features (representing distinct Dx claim dates with HTN and HTN Rx counts). This dataset was processed for modeling/ extraction of odds-ratios as described above.

### Integration of risk drivers and model predictions

Drivers for each outcome (i.e., future *atl_three_roto* status, future HTN complications, or both) in CCAE data were identified by extraction of odds-ratios using a multinomial logistic regression with 37 features (similar to those used above) listed in **Supplementary Table S3**. Separately, preceding models (predicting future *atl_three_roto* status or future HTN complications) trained on CCAE data were used to generate predictions on an HTN cohort identified from IND_DATA, overlaid for overlap/prevalence analysis of at-risk groups vs not at-risk groups, and characterized for risk stratification (based on total cost, chronic conditions burden, and utilization levels in HTN patient sub-groups).

## RESULTS

This study was performed in three stages, each representing a distinct specific aim as outlined in **Figure 1**. For the first specific aim, we focused on developing a predictive model for identifying future “challenge to treat for HTN” patients based on prior year administrative claims data and delineating “drivers” of this outcome. For this model, patients who had <u>at l</u>east <u>three</u> distinct rounds <u>o</u>f HTN treatment <u>o</u>ptions (*atl_three_roto*; see also materials and methods section) in a calendar year were defined as “challenge to treat for HTN”. Identifying these patients early is important as they have elevated future risk of HTN complications [12] and their prediction can enable timely/targeted interventions for future risk reduction within the overall HTN population. Next, to further support future risk reduction strategies, we focused on identifying “drivers” of future HTN complications in *atl_three_roto* vs *not_atl_three_roto* patients via a predictive modeling approach. Finally, we identified drivers for combinations of the above outcomes (future *atl_three_roto* status, future HTN complications, or both) and predictions from each of the models to evaluate their potential for risk stratification and management of HTN patients. Results from each of these stages are described further in this section.

**Figure 1.**
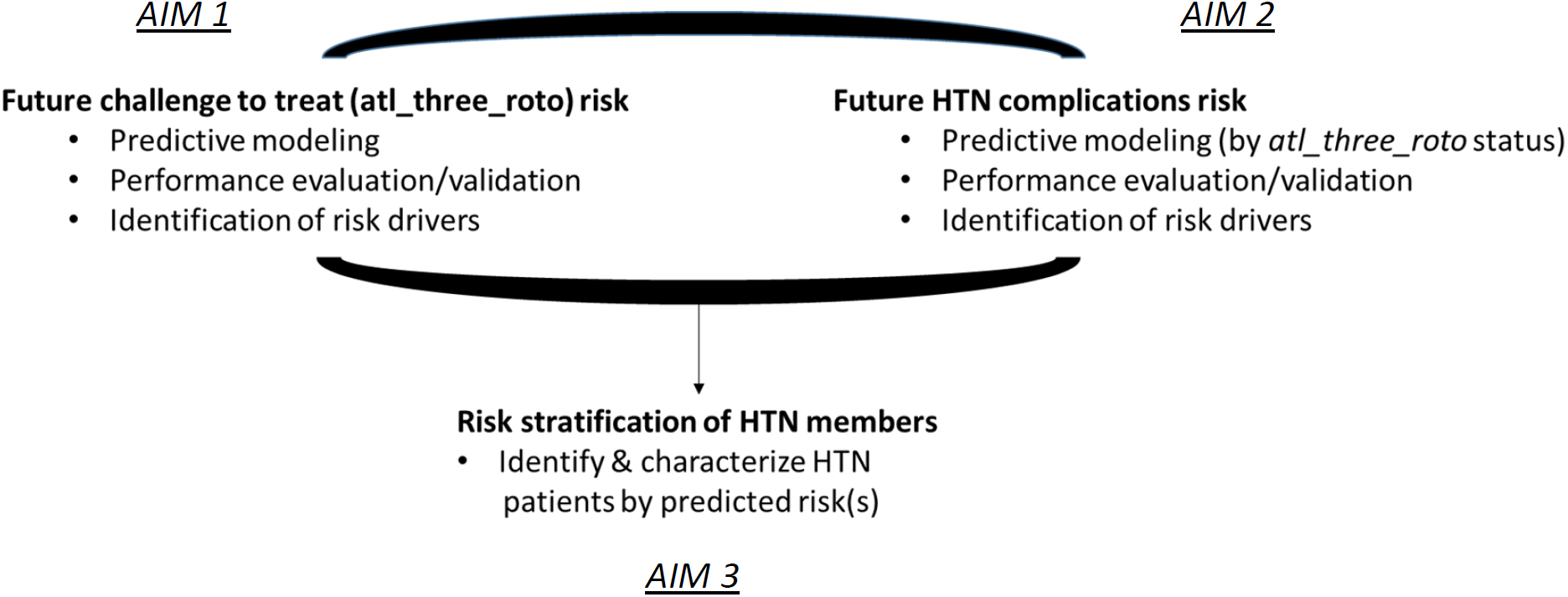
Study design/outline. Predictive models were developed for distinct risks in HTN patients in two separate stages/aims of this study and each predictive model was evaluated for performance and drivers of these risks identified/integrated. Finally, model predictions were overlaid for identification/characterization of HTN patients at risk in an external dataset (Aim 3).

### Challenge to treat for HTN predictive modeling

From our CCAE-derived study cohort of 2.47 million patients identified with HTN and continually enrolled continually through 2015-2016, 181,735 (7.3% of total) patients were labeled as “challenge to treat for HTN” in 2016 based on their *atl_three_roto* status in 2016. Previous analysis [12] had shown that weighted measures of prior-year co-morbidities (<u>C</u>harlson <u>C</u>omorbidity <u>I</u>ndex, CCI; [17]) and co-medications (<u>C</u>hronic <u>D</u>isease <u>S</u>core, CDS; [18 19]) along with selected treatment patterns (e.g., ROTO count/s i.e., rounds <u>o</u>f HTN-related distinct treatment <u>o</u>ptions) were likely associated with “challenge to treat” for HTN outcome in 2016. However, it was not clear which specific co-morbidities/co-medications were associated/predictive of this outcome and if they remained so after adjustment for other potential predictors.

Thus, we extracted from the CCAE for each member within the study cohort their specific co-morbidities, specific co-medications, age/gender information, and treatment/diagnostic pattern-related features for 2015. These were used as 45 “input” features to predict 2016 “challenge to treat” for HTN (outcome variable) status using logistic regression. A logistic regression model trained using CCAE data was evaluated across multiple metrics by cross-validation and against an independent dataset (IND_DATA; processed identically as CCAE data).

**Figure 2** shows the performance of the CCAE-trained logistic regression model using ROC-AUC curve, Precision-Recall (PR) curve, confusion matrix analyses, and additional metrics on CCAE vs IND_DATA. ROC-AUC and Precision-Recall curves were calculated using 5-fold cross-validation (*i*.*e*., 5 randomly generated test-train splits) and the confusion matrix analysis was generated using a representative test-train split. As shown in **Figure 2A left panel**, the ROC-AUC (concordance) for this predictive model is 0.84 indicating 84% probability of assigning a higher probability score to a true positive than a true negative. Due to the imbalanced nature of our dataset, ROC-AUC can overestimate the model’s predictive performance [20] and hence, we also we also generated a precision-recall curve for performance evaluation. **Figure 2A, right panel** indicates a PR-AUC of 0.39, which is ∼5.3 times better than would be expected for a “random” model (class 1/outcome prevalence of 0.073). Thus, both ROC-AUC and PR-AUC analyses show highly consistent (notice overlapping ROC/PR curves and their AUC values from each test-train split in **Figure 2A**) and substantially improved predictive performance over a baseline (random) model. From confusion matrix analysis based on a default classification threshold of 0.5 of a representative test-train split (**Figure 2B**), the model yields sensitivity/true positive rate of 73% (*i*.*e*., false negative rate of 27%) and specificity/true negative rate of 80% (*i*.*e*., false positive rate of 20%). These result in a model precision of 21.8% (∼3-fold enrichment of class 1 predictions over a random model) and balanced accuracy of 76.5%. Finally, CCAE-trained model demonstrated good generalizability across recall, precision, specificity, and balanced accuracy against an independent dataset (**Figure 2C**).

**Figure 2.**
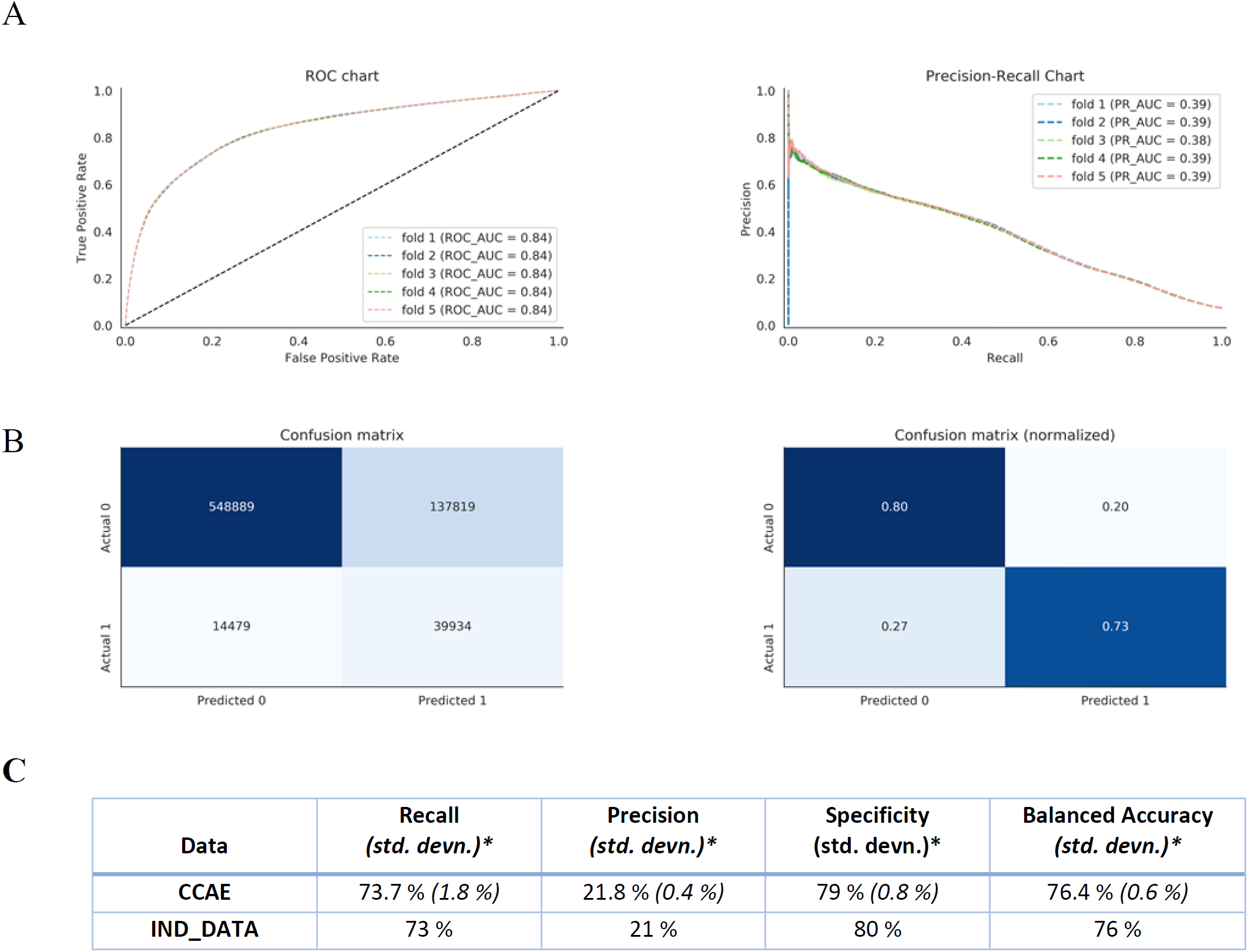
Performance evaluation of future “challenge to treat for HTN” predictive model. **A**. Receiver Operating Characteristic (ROC, left panel) and Precision-Recall (PR, right panel) curve analyses using 5-fold cross-validation (from CCAE data). **B**. Confusion matrix analyses based on default classification threshold (0.5) from a representative test-train split with raw counts (left panel) and with normalization of raw counts (right panel). **C**. Model performance metrics based on default classification threshold (0.5) on CCAE (with 5-fold cross-validation; std. devn. = standard deviation) data and IND_DATA (independent dataset).

Next, we analyzed further what features are most strongly “driving” future “challenge to treat for HTN” status. For this analysis, we extracted odds-ratios from logistic regression models trained on CCAE data. All features with at least 50% increase or decrease in odds (*i*.*e*., after rounding, odds ratio ≥ 1.5 or odds ratio ≤ 0.67) of above outcome variable are visualized in **Figure 3**. See **Supplementary Table S1** for full list of features and their associated odds.

**Figure 3.**
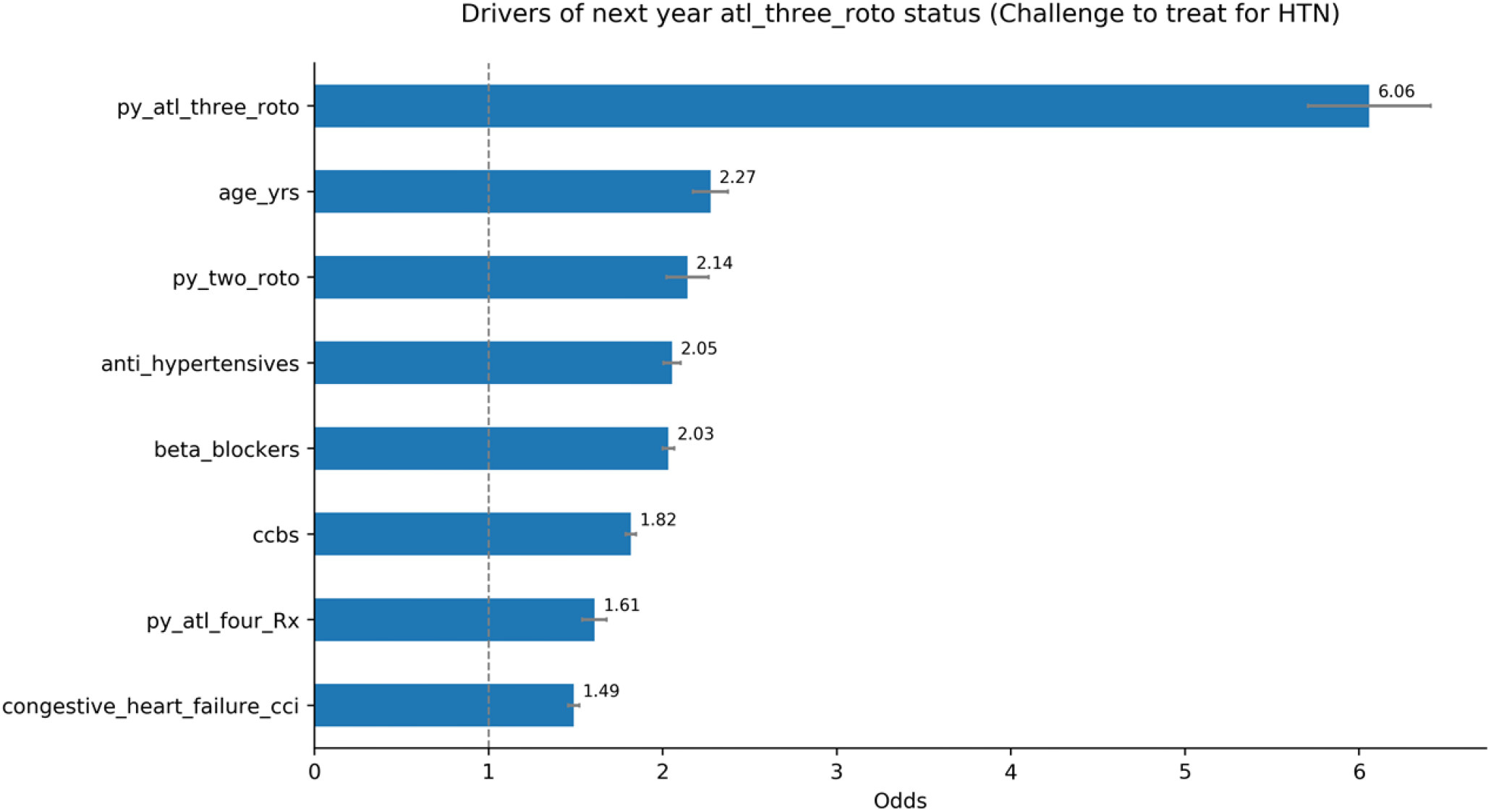
Odds-ratios for most important drivers of future (2016) *atl_three_roto* or “challenge to treat for HTN” status. All features calculated from <u>p</u>rior <u>y</u>ear (*py*, 2015) and associated with ∼50% average increase or decrease in odds are visualized. Error bars indicate 95% confidence intervals. <u>Abbreviations:</u> *py_atl_three_roto* = prior year at least three rounds of treatment options (for HTN), *py_two_roto* = prior year two rounds of treatment options (for HTN), *ccbs* = calcium channel blockers, *py_atl_four_Rx* = prior year at least four distinct Rx (medications for HTN) choices.

As shown in **Figure 3 and Supplementary Table S1**, after adjusting for other features, prior year *atl_three_roto* (*py_atl_three_roto*; <u>o</u>dds-ratio/OR = 6.1) and *two_roto* (*py_two_roto*; OR = 2.1) status are the strongest “drivers” of future *atl_three_roto* or “challenge to treat for HTN” status. Indeed, we previously found that ∼35% of *atl_three_roto* members in 2015 also had the same status in prior year (2014) indicating a “stickiness” to this higher risk group among selected HTN patients [12]. Notably, patients with ≥ 2 ROTO counts in prior year (*py_two_roto* or *py_atl_three_roto*) had higher odds of future *atl_three_roto* status than those with three (*py_three_Rx;* OR = 1.4) or more Rx choices (*py_atl_four_Rx*; OR = 1.6) in prior year – indicating that the act of changing/adding to medication choices (i.e., start of new round(s) of treatment options) rather than total number of medications in prior year is a more significant driver of future *atl_three_roto* status. Other features that increased odds of future *atl_three_roto* status include specific medication groups for HTN (anti-hypertensives/ATC class “C02” medications; OR = 2.1, beta blockers/ATC class “C07” medications; OR = 2, and CCBs *i*.*e*., <u>c</u>alcium <u>c</u>hannel <u>b</u>lockers/ATC class “C08” medications; OR = 1.8, diuretics/ATC class “C03” medications; OR = 1.3) and comorbidities (congestive heart failure; OR = 1.5, renal disease; OR = 1.3). By contrast, renin-<u>a</u>ngiotensin system acting agents (RAS agents which includes ACE inhibitors/ACEi and Angiotensin Receptor Blockers/ARBs; OR = 1.1) had much lower odds of future *atl_three_roto* status, indicating generally good tolerance/ low need to make further Rx change associated with these medications.

## Drivers of future HTN complications

In this second stage of our study, we principally sought to identify what differences, if any, there might be among *atl_three_roto* vs *not_atl_three_roto* patients with respect to factors driving future HTN complications risk. We hoped that this might better inform targeted risk reduction strategies in each of these patient sub-groups. Thus, from our CCAE study cohort, patients were partitioned into two groups based on their prior year (2015) *atl_three_roto* status and processed for predictive modeling of patients who developed next-year HTN complications followed by extraction of their “drivers” based on model-derived odds ratios.

Details of the two patient sub-groups processed further were as follows: (a) 162, 278 *atl_three_roto* patients in 2015 (43% of them developed HTN complications in 2016) and (b) ∼2.31 million *not_atl_three_roto* patients in 2015 (20.1% of them developed HTN complications in 2016). We extracted for each member within these two sub-groups, their specific comorbidities, comedications, age/gender information, and treatment /medical claim related patterns as “input features” from 2015. These were used in a logistic regression model to predict which patients in 2016 developed “any” HTN complications (see materials and methods section for additional details). **Figure 4A** demonstrates model performance metrics of the above CCAE-trained model against distinct test-train splits using CCAE data or an independent dataset (IND_DATA; processed identically as CCAE data).

**Figure 4.**
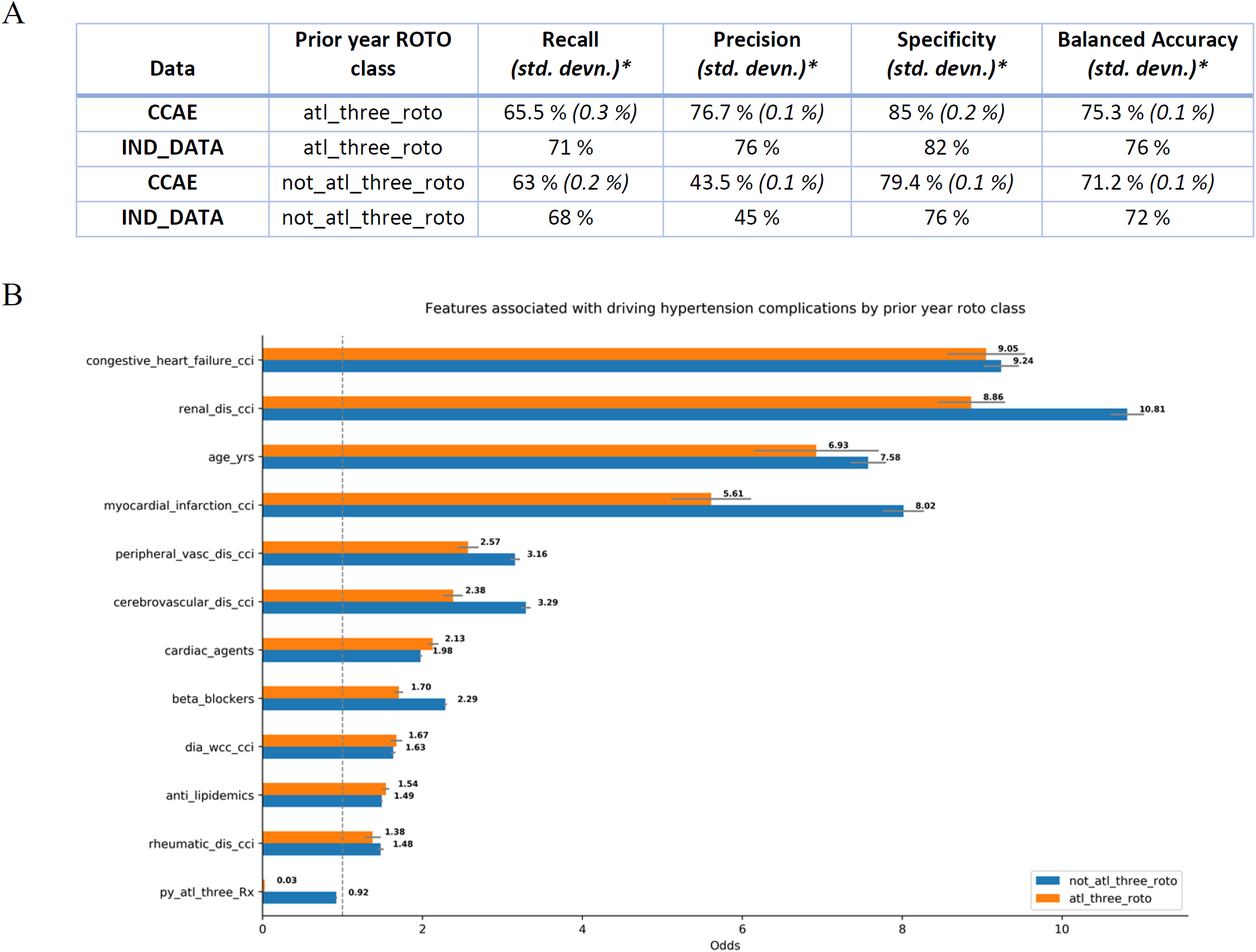
Model performance metrics and odds analysis of most important features driving future (2016) hypertension complications by prior year (2015) *atl_three_roto* status **A**. Model performance metrics for future HTN complications’ predictions based on default classification threshold (0.5) on CCAE (with 5-fold cross-validation; std. devn. = standard deviation) data and IND_DATA (Independent dataset). **B**. Odds ratios for features with at least ∼50% increase or decrease in future odds of HTN complications within *atl_three_roto* vs *not_atl_three_roto* patients. Error bars indicate 95% confidence intervals. <u>Abbreviations:</u> *renal_dis_cci* = renal disease, *peripheral_vasc_dis_cci* = peripheral vascular disease, *cerebrovascular_dis_cci* = cerebrovascular disease, *dia_wcc_cci* = diabetes with chronic complications, *rheumatic_dis_cci* = rheumatic disease, *py_atl_three_Rx* = prior year at least three distinct Rx choices for HTN.

Subsequently, we analyzed which features are most significantly associated with future (next-year) HTN complications in *atl_three_roto* vs *not_atl_three_roto* patient sub-groups. Thus, we extracted odds-ratios from above logistic regression models. All features with at least 50% increase or decrease in odds of HTN complications in either patient group were visualized for comparison (see **Figure 4B)**. See **Supplementary Table S2** for full set of features and their associated odds of future HTN complications.

Regardless of prior year *atl_three_roto* status, prior year incidence of specific comorbidities (congestive heart failure, renal disease, myocardial infarction, cerebro/peripheral vascular disease, diabetes with chronic complications, and rheumatic disease) or intake of specific medications (beta blockers, calcium channel blockers, cardiac agents/ATC class “C01” medications, and anti-lipidemics/ATC class “C10” medications) were associated with increased future odds of HTN complications.

Interestingly, for several of these “drivers” (renal disease, myocardial infarction, cerebro/peripheral vascular disease, beta blockers, prior year intake of at least 3 distinct Rx medications for HTN/ *py_atl_three_Rx*) there was a modestly subdued effect in *atl_three_roto* patients (except for *py_atl_three_Rx*). Taken together, these results indicate mostly overlapping “drivers” of future complications among *atl_three_roto* vs *not_atl_three_roto* HTN patients.

### Integration of risk drivers by future outcomes

The preceding analyses identified drivers for each of our modeling “outcomes” – occurrence of next-year *atl_three_roto*/ “challenge to treat for HTN” status and HTN complications. However, some patients may have both the preceding outcomes *vs* either of those outcomes *vs* neither of these outcomes in the following year. Thus, at least from this study’s perspective, there are 3 outcomes of interest relevant for risk stratification/management of HTN patients: (a) *ctt_for_htn_only* = Challenge to treat for HTN only (3.9% of 2.47 million patients in HTN study cohort from CCAE data), (b) *htn_comp_only* = HTN complications only (18.2% of 2.47 million HTN patients), and (c) *ctt_for_htn_&_htn_comp* = Challenge to treat for HTN and HTN complications (3.5% of 2.47 million HTN patients). To support their targeted management, we identified drivers of each of the above outcomes by a multinomial logistic regression analysis using 37 input features potentially associated with each of the above outcomes (*vs* not) in CCAE data. Subsequently, odds ratios were extracted for these features (see **Supplementary Table S3** for full results) and the strongest drivers – with at least 50% increase or decrease for any of the outcomes - are shown in **Figure 5**.

**Figure 5.**
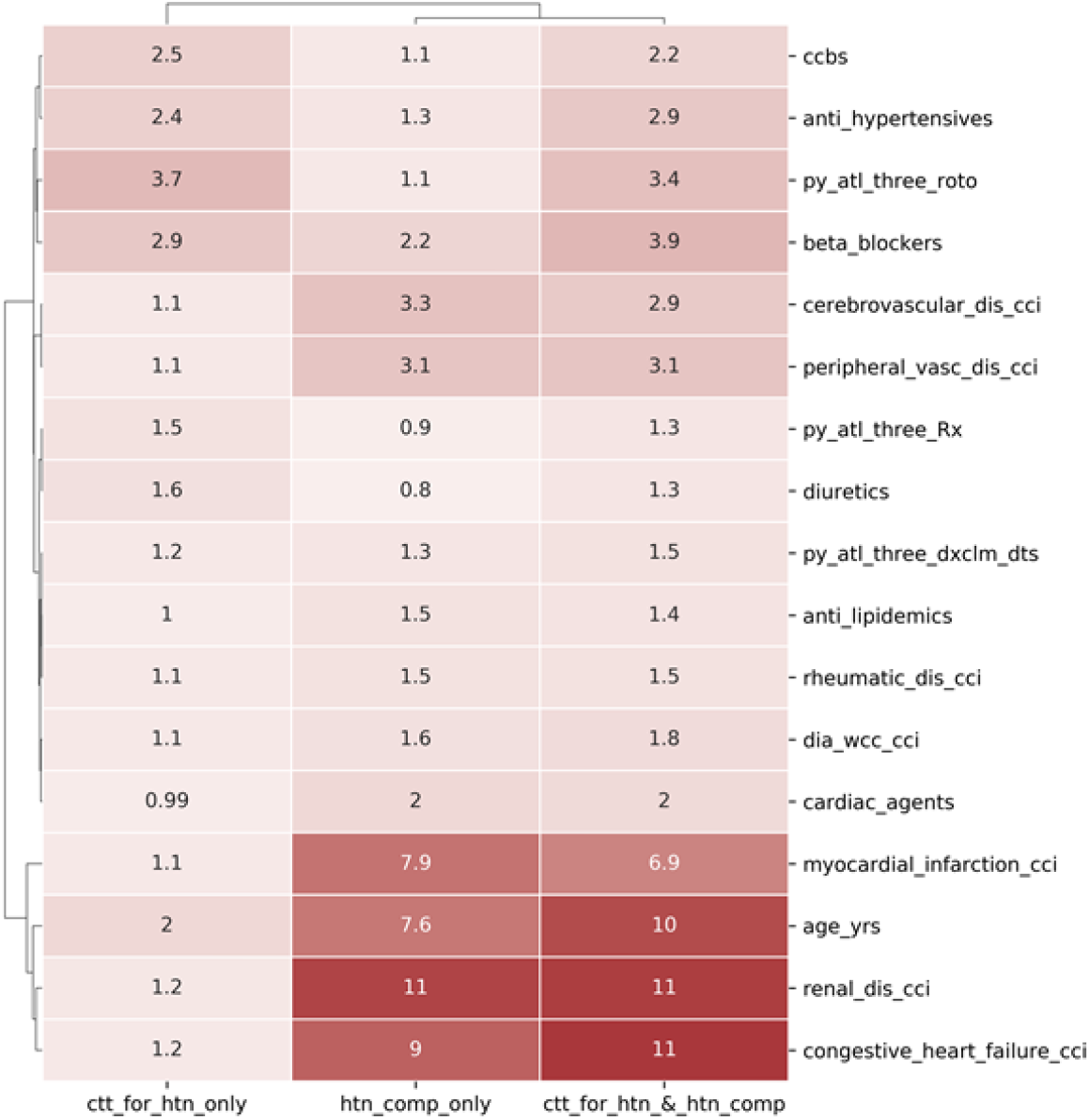
Clustered heat map analysis of selected “drivers” of future (next year) outcomes in HTN patients: *ctt_for_htn_only* = Challenge to treat for HTN only, *htn_comp_only* = HTN complications only, *ctt_for_htn_&_htn_comp* = Challenge to treat for HTN and HTN complications. Numeric value on each tile within heat map represents odds ratio associated with feature/outcome. <u>Abbreviations:</u> *ccbs* = calcium channel blockers, *py_atl_three_roto* = prior year at least three roto status, *cerebrovascular_dis_cci* = cerebrovascular disease, *peripheral_vasc_dis_cci* = peripheral vascular disease, *py_atl_three_Rx* = prior year at least three distinct Rx choices for HTN, *py_atl_three_dxclm_dts* = prior year at least three (distinct) Dx claim dates with HTN, *rheumatic_dis_cci* = rheumatic disease, *dia_wcc_cci* = diabetes with chronic complications, *renal_dis_cci* = renal disease.

As shown in **Figure 5**, there are at least three types of strongest “drivers” – those features associated with all 3 of the outcomes (*e*.*g*. beta blockers, age), those associated with either of 2 outcomes: a) *ctt_for_htn_only* and *ctt_for_htn_and_htn_complicns* (*e*.*g*. ccbs = calcium channel blockers, anti-hypertensives, *py_atl_three_roto* = prior year <u>at l</u>east <u>three r</u>ounds <u>o</u>f treatment options), or b) *htn_comp_only* and *ctt_for_htn_and_htn_complicns* (*e*.*g*. congestive heart failure, renal disease, myocardial infarction, cardiac agents, diabetes with chronic complications, rheumatic disease, and cerebro/vascular disease), and those associated strongly only with 1 outcome (*e*.*g*. diuretics with *ctt_for_htn_only)*. We hope this information can be leveraged in design of targeted care/risk management programs for HTN patient sub-groups based on their relative prevalence/importance, available resources, and magnitude of odds ratios/ relative actionability associated with these “risk-drivers”.

### Integration of model predictions for risk stratification

As a final part of our study, we sought to integrate preceding model predictions for overlap among predicted risk categories for HTN patients. Towards this aim, as a proof-of-concept analysis, we generated predictions from each of the above CCAE-trained models on an independent dataset (IND_DATA; see **Figures 2, 4A** for model performance) and characterized further our predicted sub-groups using cost, chronic disease burden and utilization levels as independent measures of risk. The results of this analysis are shown in **Figure 6**.

**Figure 6.**
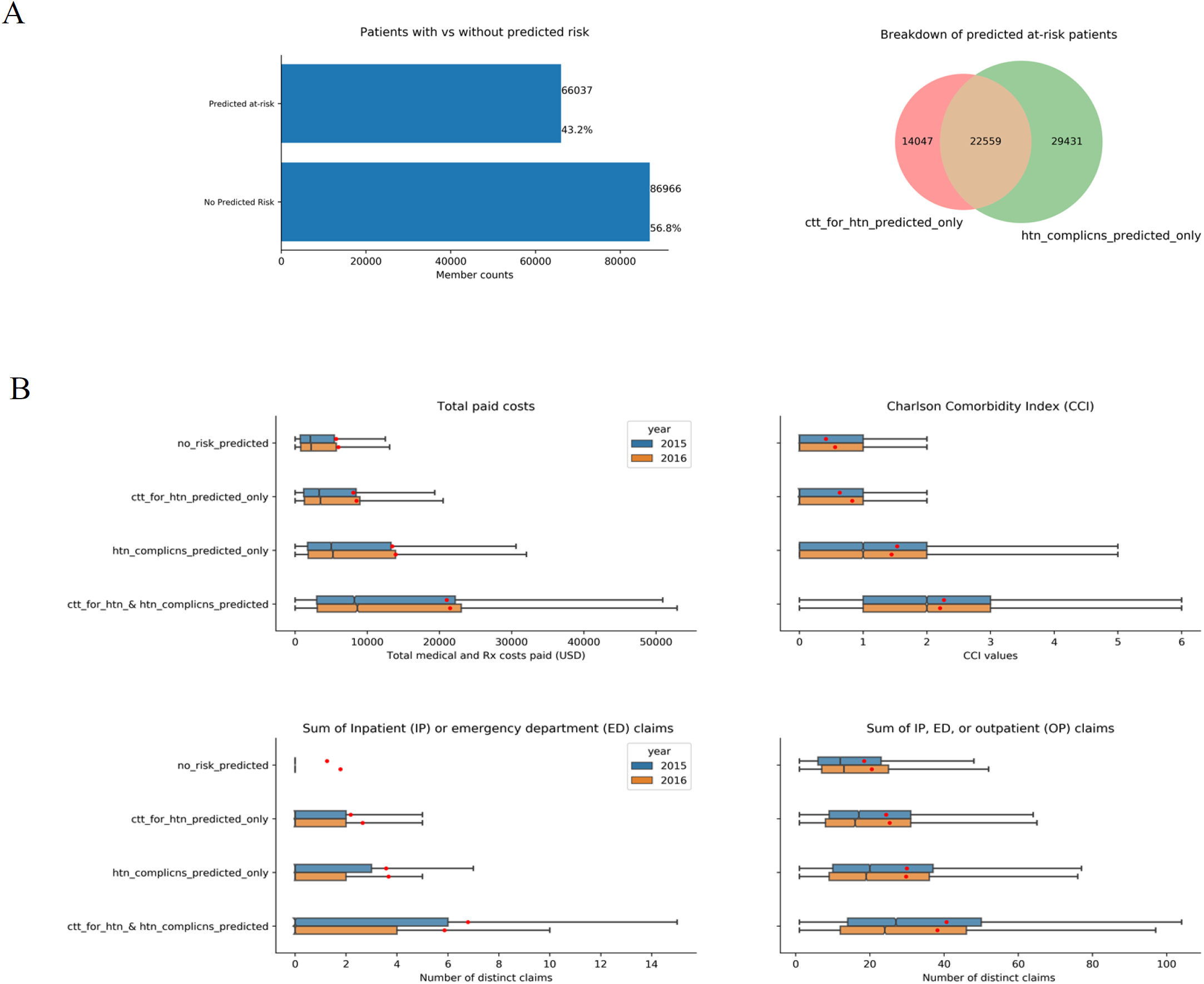
Identification and characterization of HTN patient sub-groups from independent administrative claims (IND_DATA) based on CCAE-trained model predictions for challenge to treat for HTN (*ctt_for_htn_predicted*) and future HTN complications *(htn_complicns_predi cted*) risks. **A**. Counts of identified HTN patients predicted to be “at risk” vs “not” and overlap among their predicted risk(s). **B**. Notched-box plots of total paid costs (top left panel), chronic comorbidities (CCI, top right panel), and utilization levels (bottom left panel - sum of inpatient/IP & emergency department/ED claims, bottom right panel – sum of IP, ED, outpatient claims) associated with HTN patient sub-groups for 2015 and 2016. “Notch” displays 95% confidence interval around median, red dot indicates the average and outliers are not shown.

As shown in **Figure 6A left panel**, ∼57% (86,966/153,003 total) of HTN members in IND_DATA are labeled as having “*no_risk_predicted*” as they were not predicted by our models (using default classification threshold of P = 0.5) to be at risk for either next year “challenge to treat for HTN” status or “HTN complications”. The remaining 43% of HTN patients (66,037/153,003 total) were predicted to be at risk for either/both conditions (**Figure 6B, right panel**) - 14,047 patients were predicted to be at risk for “challenge to treat for HTN” only (labeled as “*ctt_for_htn_only*”), 29,431 patients were predicted to be at risk for future HTN complications only (labeled as “*htn_complicns_predicted_only*”), and 22,559 patients were predicted to be at risk for both conditions (labeled as “*ctt_for_htn_and_htn_complicns_predicted*”). Thus, at a population level, HTN patients can be separated into 4 sub-groups based on absence/presence of each risk prediction.

As an independent measure of the associated risk with each of these sub-groups, we evaluated distributions of their total cost, chronic disease burden, and utilization levels using notched box-plot analyses (**Figure 6B**). Based on these analyses, the patient sub-groups generally in order of increasing levels of total paid cost, chronic disease burden (based on CCI), and utilization (based on distinct inpatient/emergency department claims only or distinct inpatient/emergency department/outpatient claims) levels are as following: “*no_risk_predicted”* < “*ctt_for_htn_only”* < “*htn_complicns_predicted_only”* < “*ctt_for_htn_and_htn_complicns_predicted”*. Note that the there is no clear difference in this order of increasing risk (*i*.*e*., cost/CCI/utilization) between patient sub-group in 2015 (year immediately prior to model predictions) vs 2016 (year of model predictions), indicating a consistency to these risk measures by patient sub-group year-over-year. Taken together, these analyses independently validate the utility of our predictive models for risk stratification of HTN patients.

## DISCUSSION

Among chronic diseases, HTN remains highly prevalent and challenging to control, highlighting the need for population based approaches for risk stratification/management [1]. Towards addressing this challenge, we have leveraged a predictive modeling framework using administrative claims data for early identification of two potential future risks (“challenge to treat for HTN” and “HTN related complications”) and their associated risk “drivers” among patients with hypertension. This notion is explored further in this section.

First, we developed a novel predictive model for early identification of “challenge to treat for HTN” patients. Per our definition, “challenge to treat for HTN” patients have <u>at l</u>east <u>three</u> distinct rounds <u>o</u>f HTN treatment <u>o</u>ptions (*atl_three_roto*) in a 12-month window. Identifying these patients early is important given their frequent medication changes/additions (suggesting suboptimal response to prior treatment choice/s) and their elevated risk of future/next year HTN complications. Importantly, specific HTN medications (beta blockers, calcium channel blockers, anti-hypertensives - that primarily include anti-adrenergic and arteriolar smooth muscle agents) were identified as being associated with at least 1.8-fold higher odds of future “challenge to treat for HTN” status. By contrast, first-line HTN medications (*e*.*g*. <u>R</u>enin <u>A</u>ngiotensin <u>S</u>ystem/RAS agents – which primarily includes ACEi/ARBs) were associated with significantly lower odds of future “challenge to treat for HTN” status (see **supplementary table S1**). Although there may be specific clinical conditions driving medication choices in favor of calcium channel blockers/beta blockers/anti-adrenergic agents (*e*.*g*. heart palpitations, angina, irregular heartbeat, myocardial infarction) in at least some HTN patients, our work suggests that in the absence of these conditions, alternative medication choices (*e*.*g*. RAS agents) might be useful to reduce odds of frequent medication changes/additions in future.

Next, we sought to identify “drivers” of future HTN complications in *atl_three_roto* vs remaining (*i*.*e*., *not_atl_three_roto*) HTN patients. This analysis identified mostly overlapping drivers of future HTN complications – regardless of prior year *atl_three_roto* status; these include specific comorbidities (congestive heart failure, renal disease, myocardial infarction, cerebro/peripheral vascular disease, diabetes with chronic complications, and rheumatic disease) and specific medications (beta blockers, cardiac agents, calcium channel blockers, and anti-lipidemics). Interestingly, first line therapies for HTN (diuretics and RAS agents; see **Supplementary Table S2**) were associated with either lower/even modestly protective odds of future HTN complications in both *atl_three_roto* and *not_atl_three_roto* patients. Taken together, these results suggest an HTN complications’ risk management pathway that includes (a) emphasis on best practices for management of “driver” comorbidities and (b) rationalization of medication choices based on potential risk for HTN complications (i.e., evaluating alternative choices such as diuretics/RAS agents).

An alternative approach to risk stratification/management of HTN patients might involve their initial stratification by leveraging both our predictive models to “bin” patients into sub-groups based on their predictions (*e*.*g. no_risk_predicted, ctt_for_htn_predicted_only, htn_complicns_predicted_only*, and *ctt_for_htn_and_htn_complicns_predicted)* to be followed by development of targeted interventions based on the relative prevalence of the “at-risk” sub-groups, available care management resources, magnitude of odds ratios associated with their “drivers” (see **Figure 5**), and their relative actionability.

At least a few limitations of this work deserve additional discussion. First, we focused only on predictions of future *atl_three_roto* status (based on medication changes/additions) as a way to identify/manage “challenge to treat for HTN” patients. There are other measures that also contribute to the “challenge to treat” status (e.g. social/economic determinants of health, treatment adherence, inappropriate dosages, lifestyle/behavioral patterns) that were not included in our definition for this study. Second, our analysis does not explain why *atl_three_roto* patients require multiple adjustments/changes to their HTN-Rx choices. Unfortunately, we were unable to address this limitation since we lacked access to clinical data for identified patients within this study. However, we urge potential users of our models – especially if they have access to clinical information – to further evaluate what correlation, if any, exists between ROTO counts and available clinical attributes. Third, there are other risks among HTN patients that we did not address in this work – e.g. rising cost/utilization year over year, worsening chronic disease status year over year, social/economic determinants of health, etc. Each of these likely also account for the overall challenge associated with management of HTN patients and will be addressed in future studies.

In spite of our limitations, the ability of our predictive models to identify “at-risk” HTN patients is independently validated by their elevated cost, chronic disease burden, and healthcare utilization levels. Because these risk predictions are presented along with their “drivers”, our work can support early identification and targeted management of high-risk HTN patients. Finally, we plan to evaluate in future if our predictive modeling framework – based on integrating predictions & drivers for “challenge to treat” risk with “future complications” risk - may be useful to similarly identify/manage “at-risk” patients for other chronic diseases.

## CONCLUSION

We have developed a predictive modeling based approach for risk stratification and management of HTN patients.

## Data Availability

The data used for this manuscript are commercially licensed from CCAE database and not publicly available.

## ACKNOWLEDGEMENTS

We are thankful to current/former members of Geneia LLC’s clinical team, especially, Shelley Riser, Natalie Benner, Hollie Yoder, Ronda Rogers, and Sheila Hipp, for their eager insights that guided this work.

## DATA AVAILABILITY

The data used for this manuscript are commercially licensed (CCAE, IND_DATA) and not publicly available.

---

**SUPPLEMENTARY TABLE S1.** Features and their associated odds of future (next year) “challenge to treat for HTN” status. Numeric values are rounded to two significant digits after decimal.

| **Features** | **Odds Ratio** | **+/- 95% confidence interval limits** |
| --- | --- | --- |
| **gdr** | 1.08 | 0.01 |
| **age_yrs** | 2.27 | 0.10 |
| **cerebrovascular_dis_cci** | 1.04 | 0.03 |
| **chronic_pulmonary_dis_cci** | 1.05 | 0.02 |
| **rheumatic_dis_cci** | 1.09 | 0.04 |
| **dia_wcc_cci** | 1.18 | 0.03 |
| **metas_carcinoma_cci** | 0.99 | 0.07 |
| **congestive_heart_failure_cci** | 1.49 | 0.03 |
| **myocardial_infarction_cci** | 0.97 | 0.03 |
| **peripheral_vasc_dis_cci** | 1.10 | 0.03 |
| **cancer_cci** | 1.02 | 0.03 |
| **dia_wo_cc_cci** | 1.12 | 0.02 |
| **mild_liver_dis_cci** | 1.02 | 0.02 |
| **peptic_ulcer_dis_cci** | 0.96 | 0.06 |
| **renal_dis_cci** | 1.34 | 0.03 |
| **ras_agents** | 1.09 | 0.02 |
| **anti_hypertensives** | 2.05 | 0.05 |
| **diuretics** | 1.32 | 0.02 |
| **cardiac_agents** | 1.04 | 0.02 |
| **beta_blockers** | 2.03 | 0.03 |
| **ccbs** | 1.82 | 0.03 |
| **respiratory_agents** | 1.05 | 0.01 |
| **anti_neoplastics** | 0.98 | 0.03 |
| **anti_diabetics** | 1.21 | 0.02 |
| **anti_epileptics** | 1.08 | 0.02 |
| **anti_ulcers** | 1.09 | 0.01 |
| **uric_acid_agents** | 1.16 | 0.03 |
| **anti_lipidemics** | 1.01 | 0.01 |
| **anti_migraine_agents** | 0.92 | 0.03 |
| **anti_arthritics** | 1.05 | 0.01 |
| anti_acne_agents | 1.04 | 0.02 |
| anti_glaucomics | 1.14 | 0.03 |
| glucocorticoids | 1.02 | 0.01 |
| anti_parkinson_agents | 1.08 | 0.05 |
| py_atl_three_roto | 6.06 | 0.35 |
| py_one_roto | 0.86 | 0.05 |
| py_two_roto | 2.14 | 0.12 |
| py_atl_four_Rx | 1.61 | 0.07 |
| py_three_Rx | 1.43 | 0.05 |
| py_two_Rx | 1.10 | 0.03 |
| py_atl_three_dxclm_dts | 1.13 | 0.01 |
| gdr | 1.08 | 0.01 |
| age_yrs | 2.27 | 0.10 |
| cerebrovascular_dis_cci | 1.04 | 0.03 |
| chronic_pulmonary_dis_cci | 1.05 | 0.02 |

**SUPPLEMENTARY TABLE S2.**
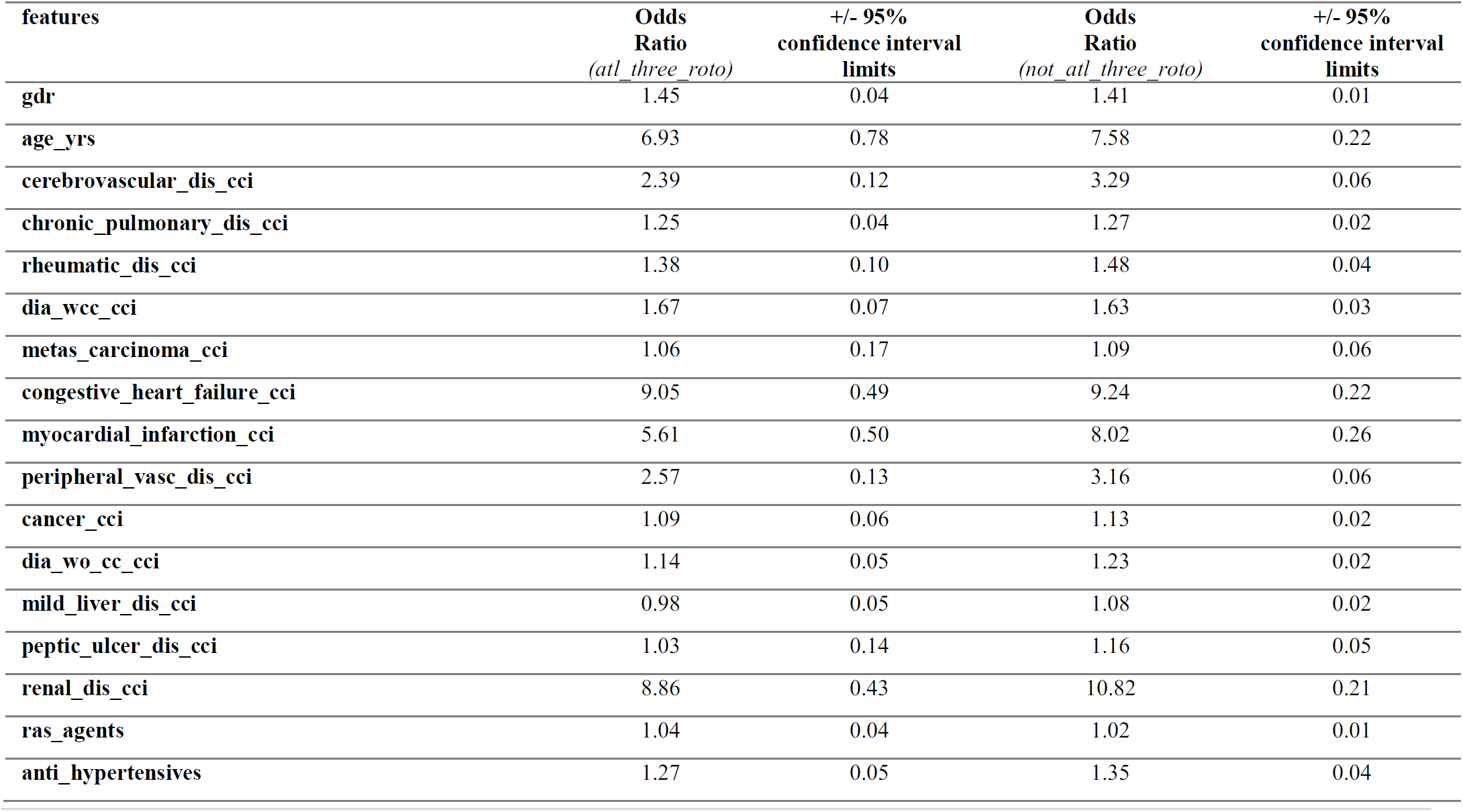
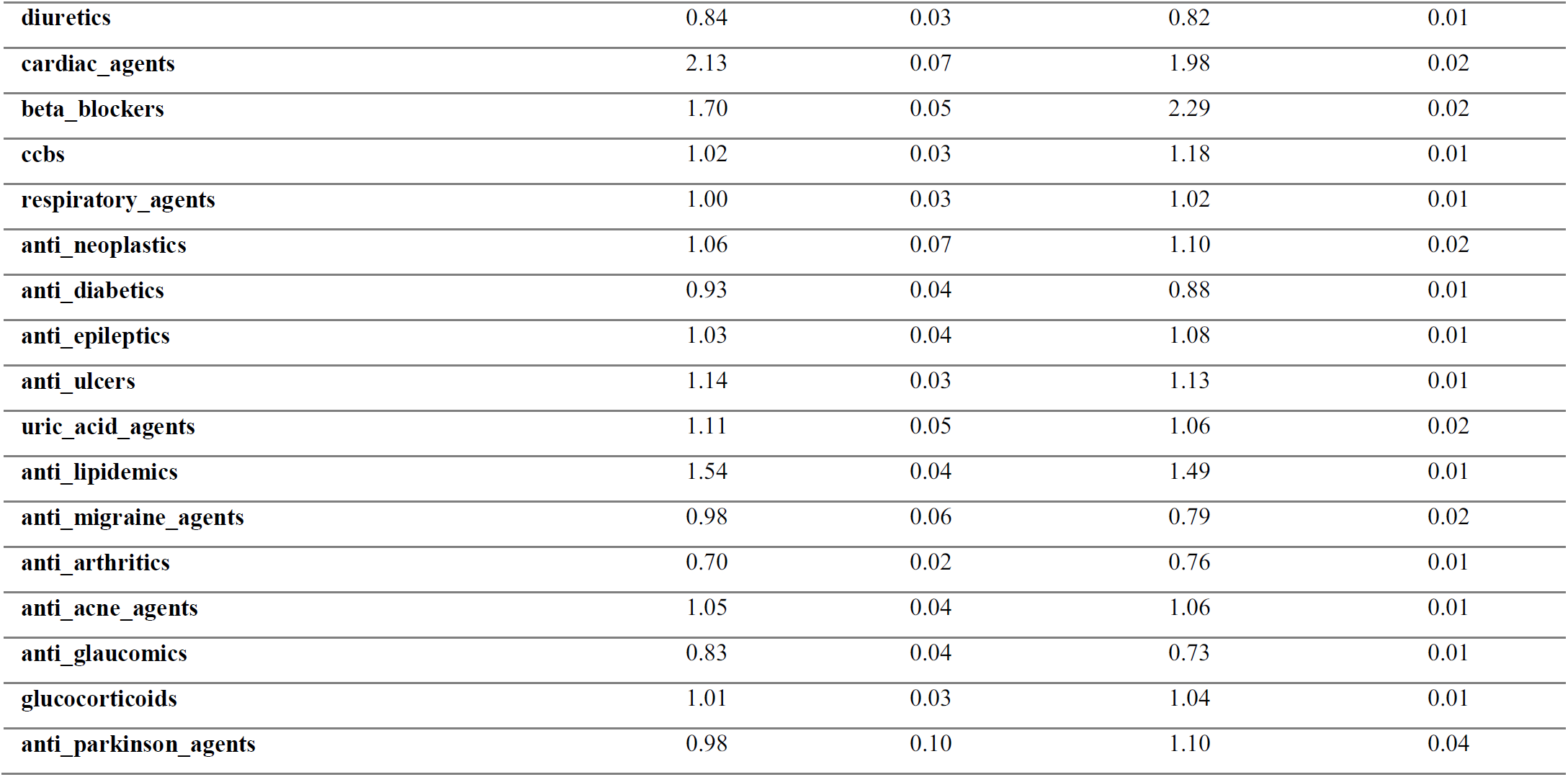
Features and their associated odds of future (next-year) HTN complications by prior year *atl_three_roto* or *not_atl_three_roto* status. Numeric values are rounded to two significant digits after decimal.

| features | Odds<br>Ratio<br>(*atl_three_roto*) | +/- 95%<br>confidence interval<br>limits | Odds<br>Ratio<br>(*not_atl_three_roto*) | +/- 95%<br>confidence interval<br>limits |
| --- | --- | --- | --- | --- |
| gdr | 1.45 | 0.04 | 1.41 | 0.01 |
| age_yrs | 6.93 | 0.78 | 7.58 | 0.22 |
| cerebrovascular_dis_cci | 2.39 | 0.12 | 3.29 | 0.06 |
| chronic_pulmonary_dis_cci | 1.25 | 0.04 | 1.27 | 0.02 |
| rheumatic_dis_cci | 1.38 | 0.10 | 1.48 | 0.04 |
| dia_wcc_cci | 1.67 | 0.07 | 1.63 | 0.03 |
| metas_carcinoma_cci | 1.06 | 0.17 | 1.09 | 0.06 |
| congestive_heart_failure_cci | 9.05 | 0.49 | 9.24 | 0.22 |
| myocardial_infarction_cci | 5.61 | 0.50 | 8.02 | 0.26 |
| peripheral_vasc_dis_cci | 2.57 | 0.13 | 3.16 | 0.06 |
| cancer_cci | 1.09 | 0.06 | 1.13 | 0.02 |
| dia_wo_cc_cci | 1.14 | 0.05 | 1.23 | 0.02 |
| mild_liver_dis_cci | 0.98 | 0.05 | 1.08 | 0.02 |
| peptic_ulcer_dis_cci | 1.03 | 0.14 | 1.16 | 0.05 |
| renal_dis_cci | 8.86 | 0.43 | 10.82 | 0.21 |
| ras_agents | 1.04 | 0.04 | 1.02 | 0.01 |
| anti_hypertensives | 1.27 | 0.05 | 1.35 | 0.04 |
| diuretics | 0.84 | 0.03 | 0.82 | 0.01 |
| cardiac_agents | 2.13 | 0.07 | 1.98 | 0.02 |
| beta_blockers | 1.70 | 0.05 | 2.29 | 0.02 |
| ccbs | 1.02 | 0.03 | 1.18 | 0.01 |
| respiratory_agents | 1.00 | 0.03 | 1.02 | 0.01 |
| anti_neoplastics | 1.06 | 0.07 | 1.10 | 0.02 |
| anti_diabetics | 0.93 | 0.04 | 0.88 | 0.01 |
| anti_epileptics | 1.03 | 0.04 | 1.08 | 0.01 |
| anti_ulcers | 1.14 | 0.03 | 1.13 | 0.01 |
| uric_acid_agents | 1.11 | 0.05 | 1.06 | 0.02 |
| anti_lipidemics | 1.54 | 0.04 | 1.49 | 0.01 |
| anti_migraine_agents | 0.98 | 0.06 | 0.79 | 0.02 |
| anti_arthritics | 0.70 | 0.02 | 0.76 | 0.01 |
| anti_acne_agents | 1.05 | 0.04 | 1.06 | 0.01 |
| anti_glaucomics | 0.83 | 0.04 | 0.73 | 0.01 |
| glucocorticoids | 1.01 | 0.03 | 1.04 | 0.01 |
| anti_parkinson_agents | 0.98 | 0.10 | 1.10 | 0.04 |

**SUPPLEMENTARY TABLE S3.**
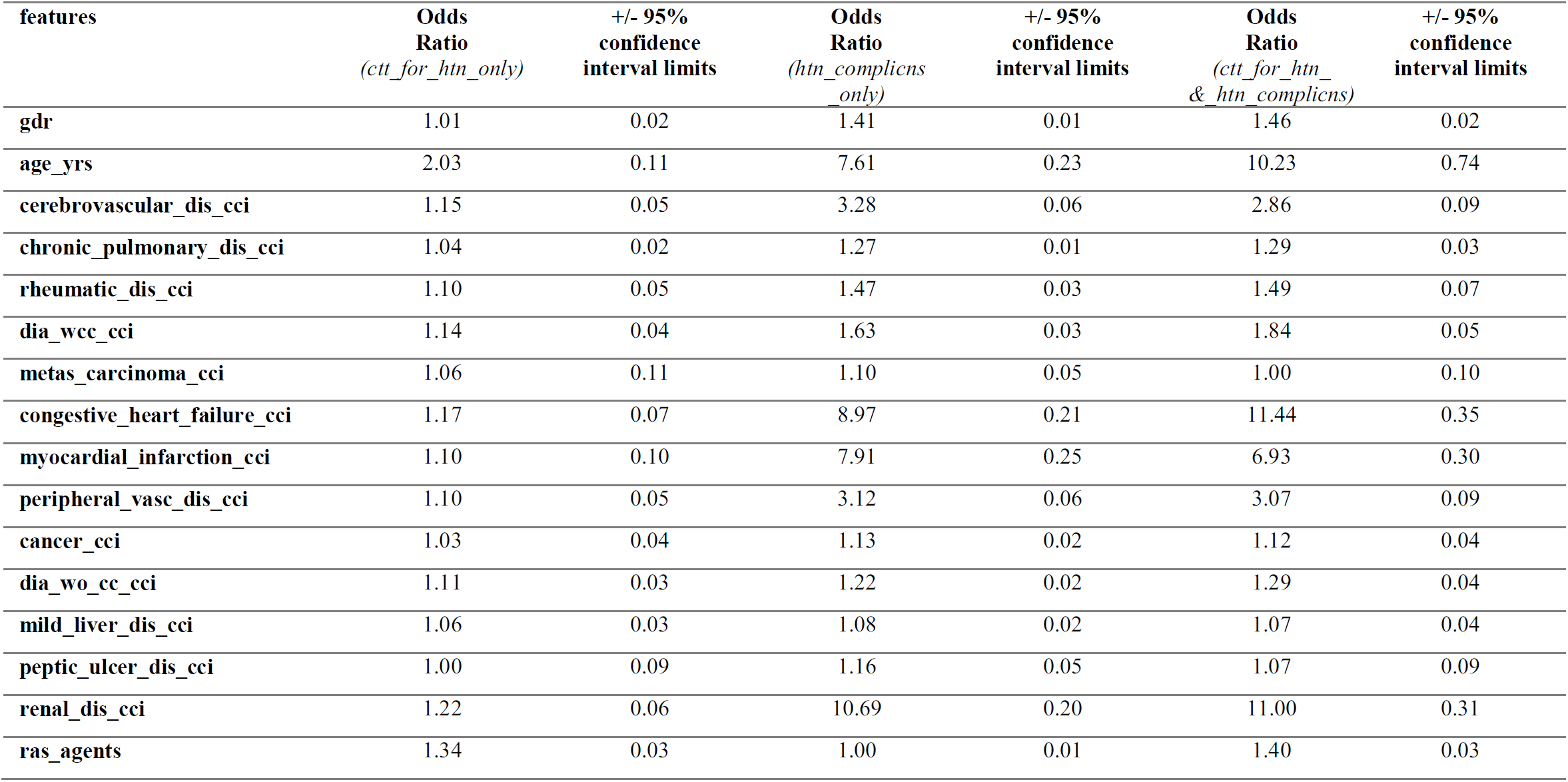
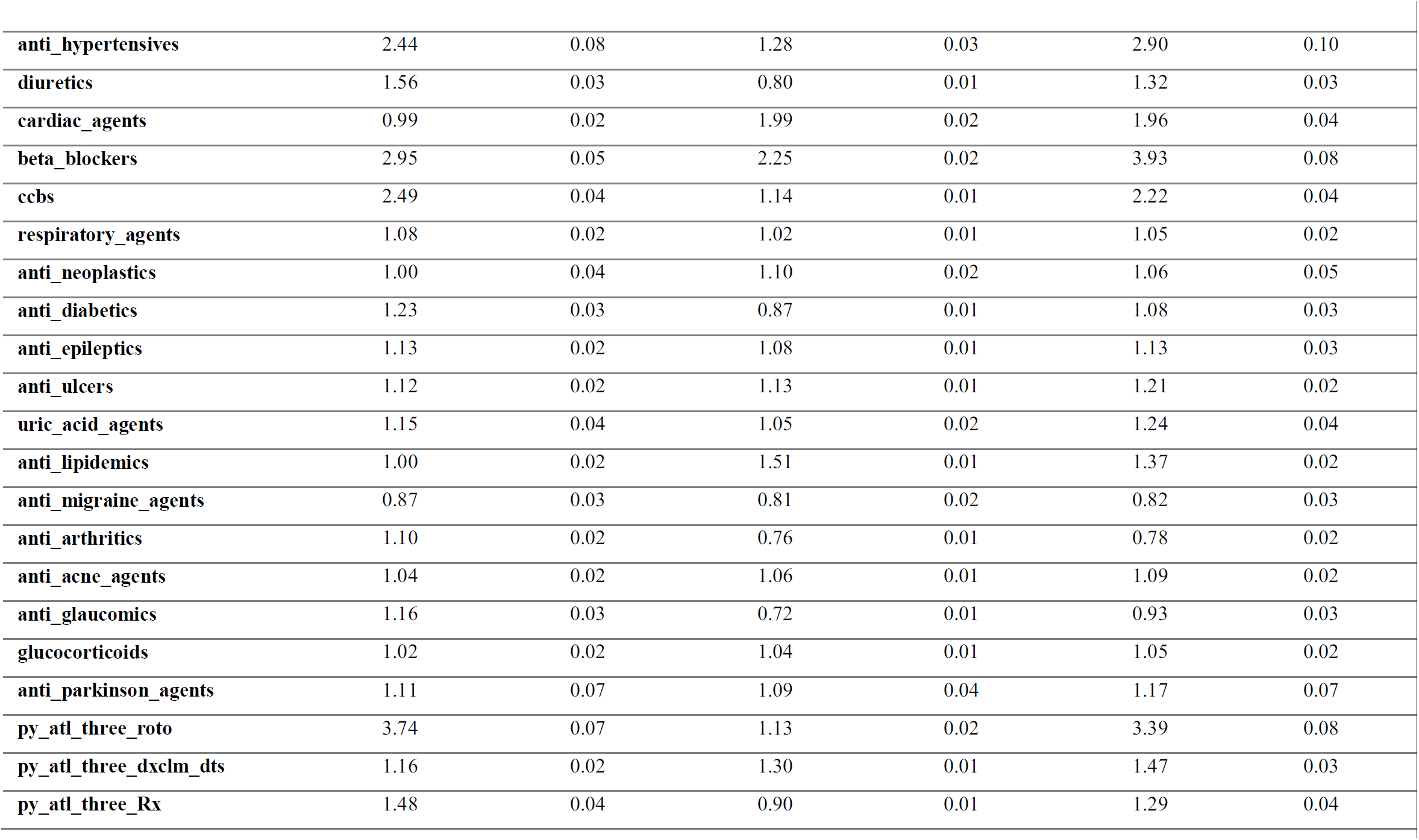
Features and their associated odds of future (next-year) risk for: challenge to treat for HTN only (*ctt_for_htn_only*), HTN complications only (*htn_complicns_only*), both outcomes (*ctt_for_htn_&_htn_complicns*). Numeric values are rounded to two significant digits after decimal.

| features | Odds Ratio<br>(*ctt_for_htn_only*) | +/- 95% confidence interval limits | Odds Ratio<br>(*htn_complicns_only*) | +/- 95% confidence interval limits | Odds Ratio<br>(*ctt_for_htn_&_htn_complicns*) | +/- 95% confidence interval limits |
| --- | --- | --- | --- | --- | --- | --- |
| gdr | 1.01 | 0.02 | 1.41 | 0.01 | 1.46 | 0.02 |
| age_yrs | 2.03 | 0.11 | 7.61 | 0.23 | 10.23 | 0.74 |
| cerebrovascular_dis_cci | 1.15 | 0.05 | 3.28 | 0.06 | 2.86 | 0.09 |
| chronic_pulmonary_dis_cci | 1.04 | 0.02 | 1.27 | 0.01 | 1.29 | 0.03 |
| rheumatic_dis_cci | 1.10 | 0.05 | 1.47 | 0.03 | 1.49 | 0.07 |
| dia_wcc_cci | 1.14 | 0.04 | 1.63 | 0.03 | 1.84 | 0.05 |
| metas_carcinoma_cci | 1.06 | 0.11 | 1.10 | 0.05 | 1.00 | 0.10 |
| congestive_heart_failure_cci | 1.17 | 0.07 | 8.97 | 0.21 | 11.44 | 0.35 |
| myocardial_infarction_cci | 1.10 | 0.10 | 7.91 | 0.25 | 6.93 | 0.30 |
| peripheral_vasc_dis_cci | 1.10 | 0.05 | 3.12 | 0.06 | 3.07 | 0.09 |
| cancer_cci | 1.03 | 0.04 | 1.13 | 0.02 | 1.12 | 0.04 |
| dia_wo_cc_cci | 1.11 | 0.03 | 1.22 | 0.02 | 1.29 | 0.04 |
| mild_liver_dis_cci | 1.06 | 0.03 | 1.08 | 0.02 | 1.07 | 0.04 |
| peptic_ulcer_dis_cci | 1.00 | 0.09 | 1.16 | 0.05 | 1.07 | 0.09 |
| renal_dis_cci | 1.22 | 0.06 | 10.69 | 0.20 | 11.00 | 0.31 |
| ras_agents | 1.34 | 0.03 | 1.00 | 0.01 | 1.40 | 0.03 |
| **anti_hypertensives** | 2.44 | 0.08 | 1.28 | 0.03 | 2.90 | 0.10 |
| **diuretics** | 1.56 | 0.03 | 0.80 | 0.01 | 1.32 | 0.03 |
| **cardiac_agents** | 0.99 | 0.02 | 1.99 | 0.02 | 1.96 | 0.04 |
| **beta_blockers** | 2.95 | 0.05 | 2.25 | 0.02 | 3.93 | 0.08 |
| **ccbs** | 2.49 | 0.04 | 1.14 | 0.01 | 2.22 | 0.04 |
| **respiratory_agents** | 1.08 | 0.02 | 1.02 | 0.01 | 1.05 | 0.02 |
| **anti_neoplastics** | 1.00 | 0.04 | 1.10 | 0.02 | 1.06 | 0.05 |
| **anti_diabetics** | 1.23 | 0.03 | 0.87 | 0.01 | 1.08 | 0.03 |
| **anti_epileptics** | 1.13 | 0.02 | 1.08 | 0.01 | 1.13 | 0.03 |
| **anti_ulcers** | 1.12 | 0.02 | 1.13 | 0.01 | 1.21 | 0.02 |
| **uric_acid_agents** | 1.15 | 0.04 | 1.05 | 0.02 | 1.24 | 0.04 |
| **anti_lipidemics** | 1.00 | 0.02 | 1.51 | 0.01 | 1.37 | 0.02 |
| **anti_migraine_agents** | 0.87 | 0.03 | 0.81 | 0.02 | 0.82 | 0.03 |
| **anti_arthritics** | 1.10 | 0.02 | 0.76 | 0.01 | 0.78 | 0.02 |
| **anti_acne_agents** | 1.04 | 0.02 | 1.06 | 0.01 | 1.09 | 0.02 |
| **anti_glaucomics** | 1.16 | 0.03 | 0.72 | 0.01 | 0.93 | 0.03 |
| **glucocorticoids** | 1.02 | 0.02 | 1.04 | 0.01 | 1.05 | 0.02 |
| **anti_parkinson_agents** | 1.11 | 0.07 | 1.09 | 0.04 | 1.17 | 0.07 |
| **py_atl_three_roto** | 3.74 | 0.07 | 1.13 | 0.02 | 3.39 | 0.08 |
| **py_atl_three_dxclm_dts** | 1.16 | 0.02 | 1.30 | 0.01 | 1.47 | 0.03 |
| **py_atl_three_Rx** | 1.48 | 0.04 | 0.90 | 0.01 | 1.29 | 0.04 |

